# The Impact of Using a Patient Health Passport on Health Outcomes among Adults with Diabetes. A Systematic Review and Meta-Analysis

**DOI:** 10.64898/2026.09.17.26363334

**Authors:** Liliana P.F. Morais, Hannah Wilson, Zena E. H. Moore, Declan Patton, Pinar Avsar

## Abstract

**Introduction:** Current evidence suggests that patient health passports may improve care for patients with chronic illnesses. This systematic review aimed to evaluate the impact of patient health passports on health outcomes in adults with diabetes.

**Methods:** A systematic review was conducted in February 2025 following PRISMA 2020 guidelines, using Ovid MEDLINE, Ovid EMBASE, EBSCO CINAHL, Web of Science, and Scopus. The review included studies of adult diabetic patients using paper or digital patient health passport as part of their care in comparison with the standard of care. Data were analysed using meta-analysis and a thematic analysis where appropriate; otherwise, the data are presented narratively. Quality appraisal was undertaken using the Cochrane Collaboration tool for assessing risk of bias and Evidence-based librarianship checklist. The certainty of the evidence was assessed using the Grading of Recommendations Assessment, Development and Evaluation.

**Results:** Eleven studies (n=14,350) were included. Seven studies were randomised controlled trials, two were retrospective in nature, one was a qualitative and one was a pre-post study. Overall, the studies are limited in design, resulting in high or unclear risk of bias across multiple domains. Results of the meta-analysis of five studies showed a statistically significant mean difference reduction in HbA1c and total cholesterol in favour of the intervention group; however, the certainty of the evidence is low or very low. The narrative analysis of the data revealed little to no improvement in body mass index, blood pressure, blood glucose, and cholesterol levels. Patients’ opinions and perceptions revealed some benefits in usability, perceived value, and in enhancing collaboration.

**Conclusion:** This review provides evidence suggesting that patient health passports may improve patient care and can inform the design of future high-quality studies. However, the low certainty of the evidence highlights the need for high-quality studies with larger sample sizes.

**Key Points:**

- Patient health passports show promise in enhancing diabetes management, however current evidence is limited by high risk of bias; future high-quality studies are required to confirm efficacy.
- Nurses are pivotal in this process as they play a central role in diabetes education, self-management support and continuity of care.
- Patient health passports could be considered as a structured tool to facilitate ongoing monitoring, reinforce adherence, enhance communication between patient and nurses/multidisciplinary team.

## 1. Introduction

Diabetes is a significant global public health issue, affecting approximately 537 million individuals worldwide, with a prevalence rate of 10.5%, and it is expected to increase by 46% by 2045^1^. Diabetes is linked with severe microvascular and macrovascular complications, reduced quality of life, higher rates of comorbidities, increased mortality risk, and increasing healthcare costs.

Similar to other chronic illnesses, diabetes management relies greatly on individuals’ self-care and routine assessments by healthcare providers^2^. Enhancing self-care skills among individuals with diabetes can help to address numerous challenges within healthcare systems^3^ and improve fundamental aspects of diabetes therapy^4^. Essential self-care behaviours include, but are not limited to, blood sugar monitoring, adherence to a healthy diet, regular physical activity, and compliance with prescribed medical treatments^4,5^.

In parallel with self-care behaviours, research strongly supports the use of diabetes care quality indicators in improving disease management, minimizing complications, and reducing treatment costs. Diabetes quality measures currently in use include glycated haemoglobin (HbA1c), blood glucose (BG), blood pressure (BP), and cholesterol levels - including low-density lipoprotein (LDL) and body mass index (BMI), which are clinical indicators of patient health status and potential risks for adverse health outcomes^6–9^. The National Glycohemoglobin Standardization Program (NGSP) and the American Diabetes Association (ADA) recommend that HbA1c testing be conducted at least twice a year^10^. In 2009, an international expert committee recognized HbA1c as a reliable indicator of chronic glycaemic levels^11, 12^, and it is now used as a gold standard index of glycaemic control^13^.

Despite recommendations, evidence suggests that almost 50% of diabetic patients do not achieve and sustain the recommended target of <7.0% for HbA1c, and only 14.3% are at target goals for HbA1c, low-density lipoprotein cholesterol, and BP^14^. Evidence suggests that improving cholesterol, hypertension, and BG levels can significantly reduce the risk of adverse clinical outcomes and treatment costs^15^. In Europe, there have been positive trends in achieving treatment targets for meeting all three key health measures: HbA1c, blood pressure and LDL cholesterol^16^.

Diabetic care demands a complex approach. This should include periodic assessments of metabolic control and complications, a collaboratively developed and continuously updated diabetes care plan, and access to personalized, multidisciplinary care^17^. There has been a growing consensus that incorporating the multidisciplinary team (MDT) approach to diabetes care provides an integrated, high-quality, and patient-centred care^17^. However, despite these benefits, significant gaps are found in the communication and coordination between providers and patients. The high proportion of diabetes patients reporting issues with care coordination highlights the urgent need for targeted quality improvement efforts in this area^18^.

Effective communication is crucial for healthcare services as it influences the quality of healthcare delivery, affects patient health and satisfaction and ultimately benefits both patients and providers^19^. Evidence suggests that communication and planning aids, including paper or digital health passports, provide a personalised health information tool^20^. A patient health passport (PHP) is a tool that provides individuals with personalised health information through mobile devices or physical documents containing their medical records^20, 21^. The patient health passport paper (PHPp) format enhances person-centred care experience^22^, improves health knowledge and patient’s attitude^23^, improves communication across a range of service providers and patients^24, 25,26^, enhance continuity of care^27^, encourage self-management^24^, engagement^28^ and autonomy^27, 29^, strengthening the therapeutic alliance^29^, building integrated care^29^ and reduces health inequalities^30^.

A digital PHP, also referred to as a personalised health record (PHR), plays a role in ensuring continuity of care, and along with advancements in technology and patient capabilities, has facilitated the transition from PHPp to digital/electronic records^20, 21^. It plays a key role in improving information management and supports more informed medical decision-making^31^. Collective evidence has shown high acceptability^32^, increased usage of medical services among patients^20^, enhanced patients’ knowledge and autonomy^33, 34^, empowerment^34–36^, improved quality of care^20^, prevented adverse effects^35^, enhanced healthcare delivery service^20^ and could offer a scalable, cost-effective approach to enhancing care for patients with chronic physical and mental illnesses^20^. PHRs could play a critical role in supporting patients with chronic conditions like diabetes by improving continuity of care, facilitating the exchange of information between patients and physicians, and ultimately enhancing quality care outcomes^37, 38^. Critically, some studies have addressed limited aspects of PHRs, including communication challenges, the impact on doctor-patient relationships^39^, the invisible work required of patients^40^ and the practical implications for patients’ lives^40^. Additionally, there is ongoing debate about the lack of sufficient research and systematic reviews on the use of PHR among patients^3, 41^. Therefore, this systematic review (SR) aims to address this gap by systematically reviewing the impact of using a PHP on health outcomes among adults with diabetes.

## 2. Aim

The aim of this systematic review was to evaluate the impact of using paper and digital PHP on health outcomes among adults with diabetes.

## 3. Materials and Methods

### 3.1 Design

A systematic review of the published literature was conducted following the recommendations and guidelines outlined in the Cochrane Handbook for Systematic Reviews of Interventions^42^. Additionally, the principles of the Preferred Reporting Items for Systematic Reviews and Meta-Analyses (PRISMA) were adhered to^43^. The protocol was registered by the National Institute for Health Research (NIHR) in the PROSPERO international prospective register of systematic reviews. The study question was developed using a structured approach referred to as PICO^44^ (population, intervention, comparator and outcomes). The specific elements of PICO were as follows:

• **P (Population):** Adult diabetic patients.

• **I (Intervention):** PHPp and PHR.

• **C (Comparison):** Standard care without a PHPp or PHR.

• **O (Outcome):** Primary Outcome: Diabetes health outcomes - HbA1c, BMI, BP, Cholesterol levels (total and LDL); Secondary Outcomes: Patients’ opinions and perceptions of the PHPp or PHR.

Thus, the question was: What is the impact of using a patient health passport on health outcomes among adults with diabetes?

### 3.2 Search Methods

The following databases were searched in February 2025: Ovid MEDLINE, Ovid EMBASE, EBSCO CINAHL, Web of Science and Scopus, with no search date limitations applied. Based on the formulated research question, two main keywords were identified: *Patient health passport* and *Diabetes*. To expand these keywords, the authors explored search terms by reviewing the abstracts of papers known to be relevant from the outset and drawing on their knowledge from their field/discipline/area of study to compile a collection of terms relevant for constructing the search strategy. Additionally, they considered keyword suggestions by seeking expert opinions. The finalised list of keywords searched in each database is presented Table 1. Following the database searches, two independent authors identified the selected papers by screening them manually. Article titles were evaluated and the abstracts of studies identified through the search strategy were reviewed for eligibility based on predefined inclusion and exclusion criteria. Then, full-text versions of studies deemed potentially relevant were retrieved, and two authors independently assessed these against the inclusion criteria. Studies of both quantitative and qualitative designs were included because the review aimed to evaluate both clinical outcomes and patient experiences associated with patient health passport use. Quantitative evidence informed effectiveness outcomes, while qualitative evidence contributed to understanding usability, acceptability and patient perceptions. Consensus between the two authors in relation to the studies and the data to be included was obtained through a discussion when discrepancies were identified. A PRISMA flow chart^43^ was created to provide a visual display of literature flow and the final corpus of citations included.

**Table 1.** Search strategy used, including database and Keywords.

| No. | Query | Last Run Via | Results |
| --- | --- | --- | --- |
| 1 | ((patient* or health or care or hospital) adj2 passport*) or ((patient or personal) adj2 "health record*") or ("patient held communication tool*" or "patient centered care document*" or "patient-held medical history document*" or "communication passport*")).mp. | Ovid Medline | 3780 |
| 2 | exp Diabetes Mellitus/ or (diabetes or diabetic).mp. | Ovid Medline | 920052 |
| 3 | 1 and 2 | Ovid Medline | 265 |
| No. | Query | Last Run Via | Results |
| #1 | ((patient* OR health OR care OR hospital) NEAR/2 passport*) OR ((patient OR personal) NEAR/2 'health record*') OR 'patient held communication tool*' OR 'patient centered care document*' OR 'patient-held medical history document*' OR 'communication passport*' | Embase | 3,669 |
| #2 | 'diabetes mellitus'/exp OR diabetes OR diabetic | Embase | 1,785,499 |
| #3 | #1 AND #2 | Embase | 498 |
| No. | Query | Last Run Via | Results |
| S1 | ((patient* or health or care or hospital) N2 passport*) OR ((patient OR personal) N2 "health record*") OR "patient held communication tool*" or "patient centered care document*" or "patient-held medical history document*" or "communication passport*" | Ebsco Cinahl | 2,327 |
| S2 | (MH "Diabetes Mellitus+") OR diabetes or diabetic | Ebsco Cinahl | 298,945 |
| S3 | S1 AND S2 | Ebsco Cinahl | 213 |
| No. | Query | Last Run Via | Results |
| #1 | TS=((patient* or health or care or hospital) NEAR/2 passport* ) OR ((patient OR personal) NEAR/2 "health record*") OR "patient held communication tool*" or "patient centered care document*" or "patient-held medical history document*" or "communication passport*") | Web of Science | 5,770 |
| #2 | TS=( diabetes or diabetic ) | Web of Science | 1,023,300 |
| #5 | #1 AND #2 | Web of Science | 515 |
| No. | Query | Last Run Via | Results |
| 1 | TITLE-ABS-KEY ( ( ( patient* OR health OR care OR hospital ) W/2 passport* ) OR ( ( patient OR personal ) W/2 "health record*" ) OR "patient held communication tool*" OR "patient centered care document*" OR "patient-held medical history document*" OR "communication passport*" ) | Scopus | 9,036 |
| 2 | TITLE-ABS-KEY ( diabetes OR diabetic ) OR INDEXTERMS ( "Diabetes Mellitus" ) | Scopus | 1,386,888 |
| 3 | 1 AND 2 | Scopus | 761 |

To identify further published, unpublished and ongoing studies, this systematic review also:

### • Searched conference proceedings, research reports and dissertations

• Scanned reference lists of all identified studies and reviews to assess for further relevant citations;

• Performed a manual search of relevant literature to enhance the capture of relevant and unique literature.

### 3.3 Inclusion and Exclusion Criteria

#### Inclusion criteria

§ Original peer-reviewed studies published in English.

§ Diabetic patients ≥18 years old.

§ Adult diabetic patients using a PHPp or PHR as part of their care.

§ Studies of both qualitative and quantitative research designs.

#### Exclusion criteria

§ Case study/ case series.

§ Adult non-diabetic patients using a PHPp or PHR as part of their care.

§ Adult diabetic patients not using a PHPp or PHR as part of their care.

§ Adult diabetic patients using mobile health apps as part of their care

### 3.4 Quality Appraisal

The methodological quality of the qualitative included articles was assessed using the Evidence-based Librarianship (EBL) Checklist ^45^. This quality appraisal tool assesses the validity, applicability, and appropriateness of the study, based on four main steps of the research process: Population; Data collection; Study design; Results. According to this checklist, if the overall validity of the study (Yes/Total) is ≥75% or (No + Unclear/Total) is≤25% then the study is valid^45^. The risk of bias (RoB) of the included RCT’s studies was assessed using the Cochrane Collaboration risk of bias tool^42^. This tool addresses six specific domains, namely sequence generation, allocation concealment, blinding, incomplete outcome data, selective outcome reporting and other bias. Two review authors independently assessed the RoB for each study and resolved any disagreements through consensus. The results of the RoB assessment were incorporated into an overall grading of the evidence related to each of the main outcomes using GRADE (Grades of Recommendation, Assessment, Development and Evaluation)^46^. The certainty of the body of evidence was then assessed against five principal domains: 1. Limitations in design and implementation; 2. Indirectness of evidence or generalisability of findings; 3. Inconsistency of results, for example, unexplained heterogeneity and inconsistent findings; 4. Imprecision of results was confidence intervals were wide; and 5. Publication bias^42^.

### 3.5 Data Collection

Data from the included articles were extracted and entered into a pre-designed table using the following headings: Author and date of the study, setting, geographic location, sample size and characteristics, design, intervention, comparison, primary outcomes: Diabetes health outcomes (HbA1c, BMI, BP, Cholesterol levels); secondary outcomes: Patients’ opinions and perceptions of the PHPp or PHR and limitations.

### 3.6 Synthesis

Firstly, the data were narratively summarised using percentages (number) and average (standard deviation (SD)) or median (range), which were appropriate, giving an overview of the study design, geographical location, study settings, sample sizes, intervention and comparison. Then, where appropriate, data were analysed using meta-analysis, statistical synthesis, thematic analysis and a narrative analysis. Meta-analysis statistical synthesis was undertaken using RevMan^47^. Relative risks (RRs) and 95% confidence intervals (CI) were calculated for dichotomous outcomes (PHP and Non-PHP) and results were presented as mean difference.

For the thematic analysis of the patients’ opinions and perceptions of the PHPp or PHR, both the results and interpretations sections of papers were assessed, following the approach of Thomas and Harden^48^. The analysis was conducted manually to maintain close engagement with the data. Thematic synthesis involved three overlapping stages: (i) free line-by-line coding of the meaningful data segments; (ii) the consolidation of these ‘free codes’ into related areas to develop descriptive clusters, which were used to (iii) generate analytical themes, through reflection and discussion with all authors. These analytical themes go beyond the primary studies and directly address the aims of the review.

#### Assessment of Heterogeneity

Results of comparable trials were pooled using either a fixed-effect model or random effects model, depending on heterogeneity, which was investigated using the *I*^2^ statistic. Heterogeneity was assessed using the *I^2^* test^49^. Where there was evidence of substantial heterogeneity (*I*^2^ > 50%), we used a random effects model in the meta-analysis. In the absence of significant heterogeneity, a fixed effects model was employed in the meta-analysis^49^.

## 4. Results

### 4.1 Results of the Search

As shown in Figure 1, following reviews of titles and abstracts from a total of 2252 citations, 1145 were found to be duplicates and removed. After screening title and abstracts of 1107 citations, 1075 studies were subsequently excluded. Following a review of the full texts of the remaining 32 studies, a further 21 were excluded: 15 studies^50–64^ had different interventions; four had different outcomes^65–68^; one study^69^ had a different comparator and one study^70^ was duplicated with different publishing dates. This left 11 studies^3, 71–80^ that met the inclusion criteria, and these form the basis of this SR.

**Figure 1:**
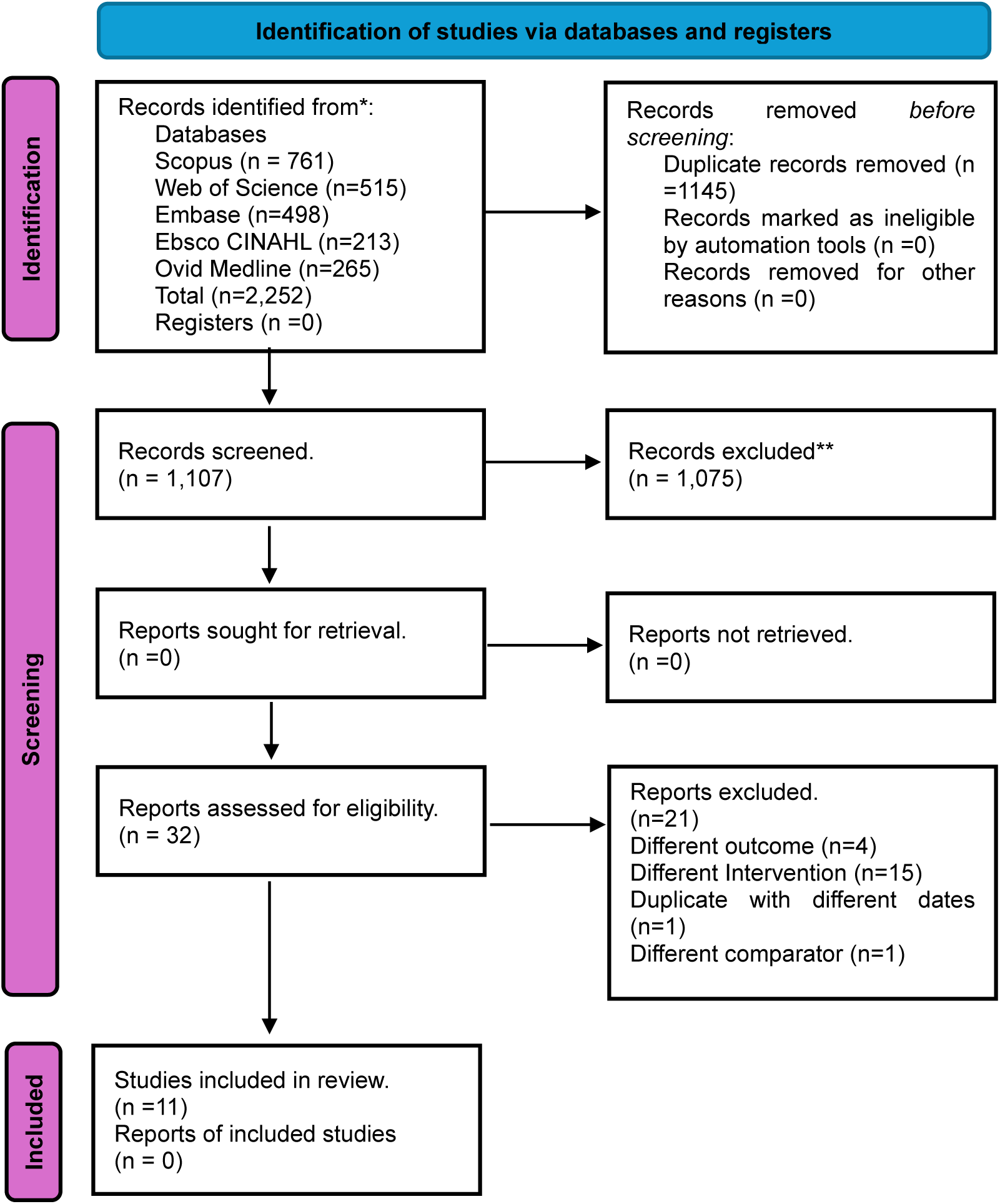
**Prisma flow diagram of the selected studies**

### 4.2 Description of the Included Studies

#### 4.2.1 Study Designs

The studies were published between 2013 and 2023. Seven studies^3, 71–73, 75, 76, 79^ were RCTs, two^78, 80^ were retrospective in nature, one was a qualitative study^74^, and one was a pre-post study^77^.

#### 4.2.2 Geographical Location

Five^74–77, 80^ of the studies were conducted in the United States of America (USA), three^71–73^ were conducted in the Netherlands, one in Japan^78^ and another in Iran.

#### 4.2.3 Study Settings

The most common study setting was the community (*n*=7, 64%), including general practice^73, 79^, primary care^76, 80^, and one endocrinology clinic^3^. One study did not state the specific setting^77^.

#### 4.2.4 Sample Size

The sample sizes ranged from 50 participants^77^ to 10746 participants^80^, with an average size of 1304.6 (SD:± 3002.5) participants, from a total of 14,350 participants.

#### 4.2.5 Intervention and Comparison

Seven studies^3, 74–78, 80^ compared PHR versus usual care; 3 studies^71, 73, 79^ compared PHPs versus usual care and 1 study^72^ compared PHPs versus usual and versus professionally directed group.

### 4.3 Follow-up

The follow-up time in the studies was a median of 6 months, which ranged from three months^77^ to twelve months^71, 72, 76, 79, 80^.

### 4.4. Results for the Risk of Bias Assessment

Figure 2 provides an overview of the RoB assessment of 10 of the included studies^3, 71, 72, 74–80^. One study^74^ was not included in this assessment, as it was a qualitative study. As can be seen, all studies had areas of high risk of bias, with the most notable being a lack of blinding of participants, personnel and outcome assessors. The exception was Simmons^79^, who addressed the blinding of participants through use of identical covers on the passport and control booklets. A further issue of concern was the lack of use of robust methods of randomisation^72, 73, 75, 77, 78, 80^ and a high or unclear risk of bias in terms of allocation concealment in all studies, except for Dijkstra^71^. Furthermore, there was a high or unclear risk of attrition bias in all studies, except for Grant^76^.

**Figure 2:**
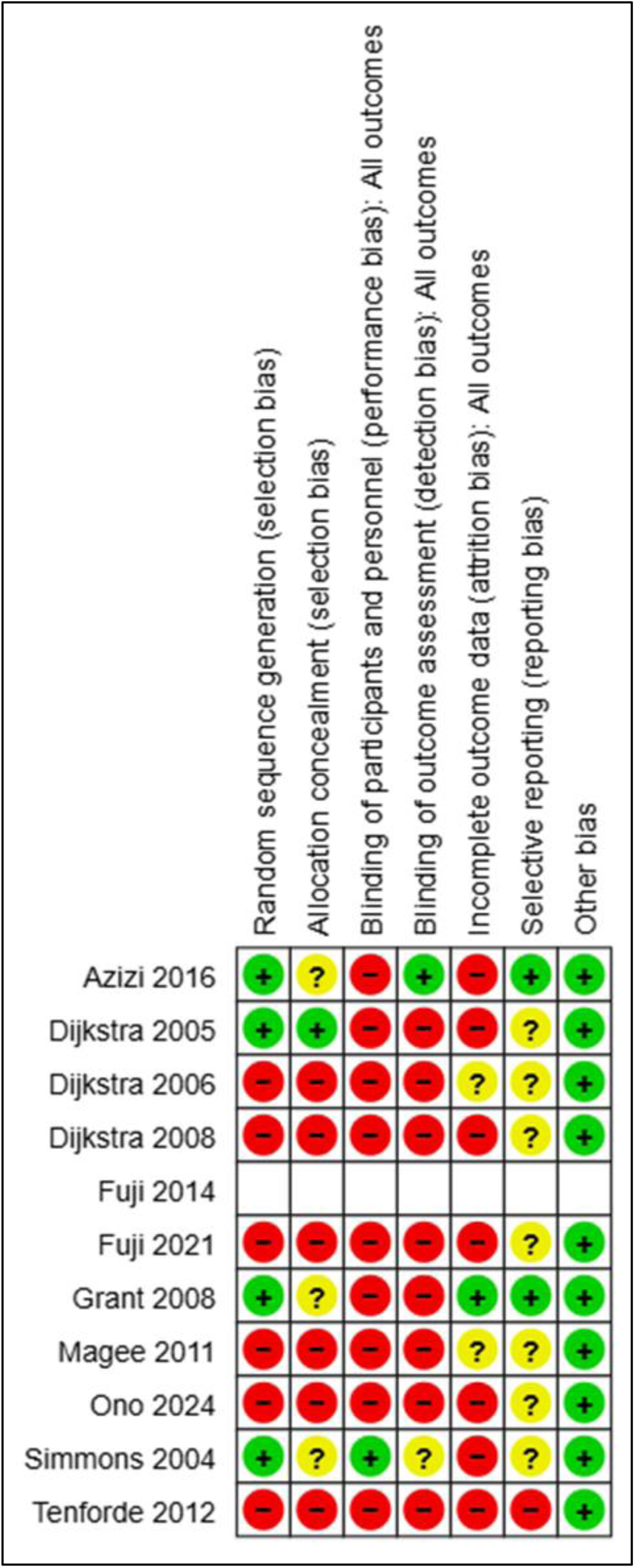
**Risk of bias summary of the included studies**

The EBL appraisal checklist was employed to critically assess the methodological rigour of the included qualitative study^74^ and was deemed not valid. In the domain of population, the study scored <75% in validity. When considering the population domain, the key areas of concern are related to the fact that it was unclear if the population sample size was large enough to be representative and to give precise estimates. The study did not state if consent was obtained. For the data collection domain, the study scored <75% in validity. The primary issues of concern were associated with the unclarity if instrument was bias reduced or validated and whether those involved in data collection were also involved in the delivery of service. In the study design domain, it scored 80% (valid), however it was unclear if ethics approval was granted. Additionally, in the results domain scored 50% for validity. Key issues related to uncertainty if studies have external validity which was found in all the studies and if confounding variables were accounted for. Evener, it did not clearly provide suggestions for further areas to research.

### 4.5 Outcomes

#### 4.5.1 Summary of the outcomes measured in the Included Studies

As can be seen on Table 2, for the primary outcome, all eleven studies^3, 71–80^ (100%)analysed HbA1c; eight studies^3, 71–73, 75–77, 79, 80^ (73%) analysed BP; ten studies^3, 71–73, 75–80^ (91%) analyse cholesterol; three studies analysed BG^74, 77, 78^ (27%) and five studies^72, 75, 78–80^ (45%) analysed BMI. Five studies^3, 73–75, 79^ (45%) analysed patients’ opinions and perceptions (see Table 3).

**Table 2.**
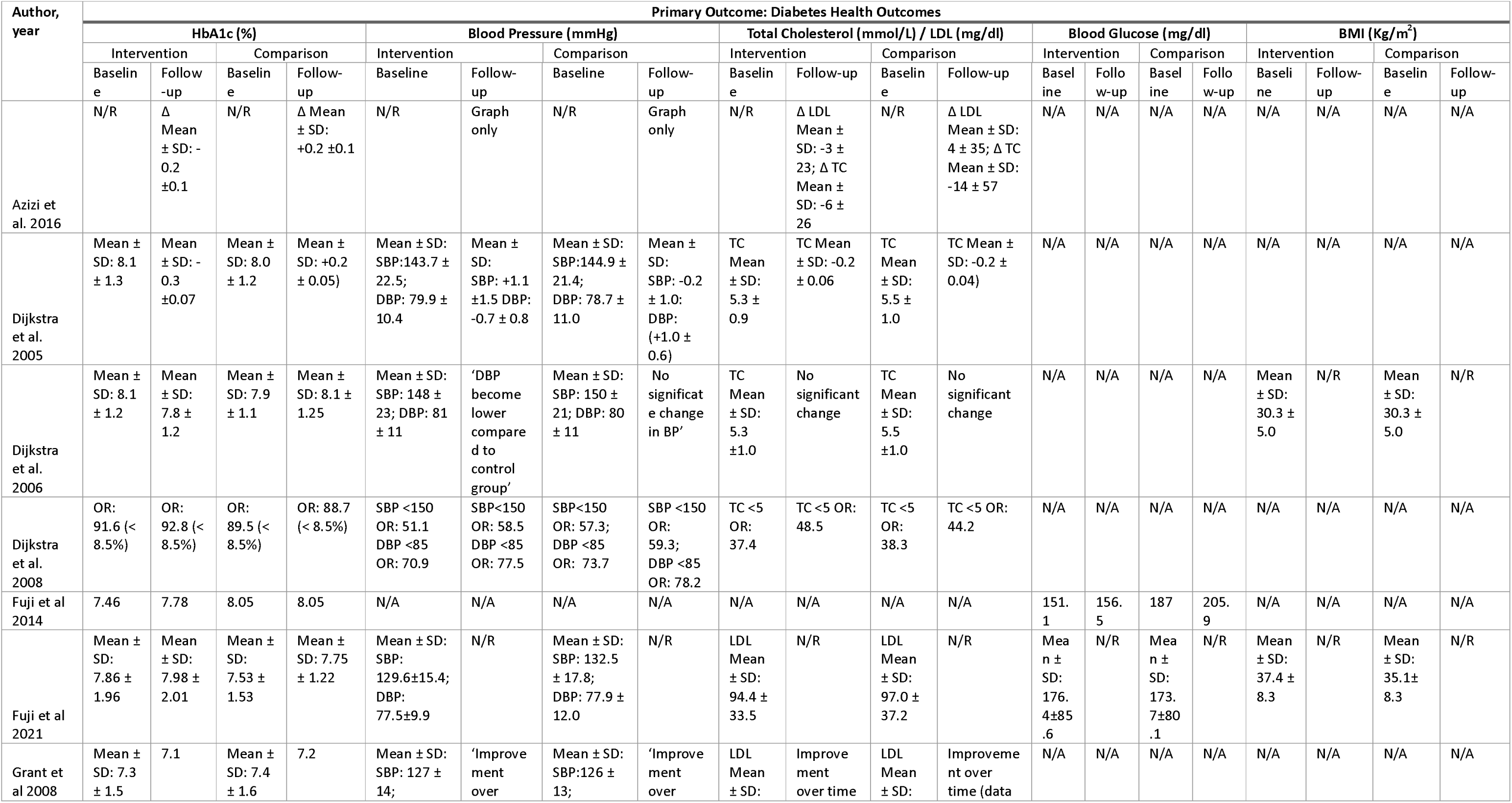
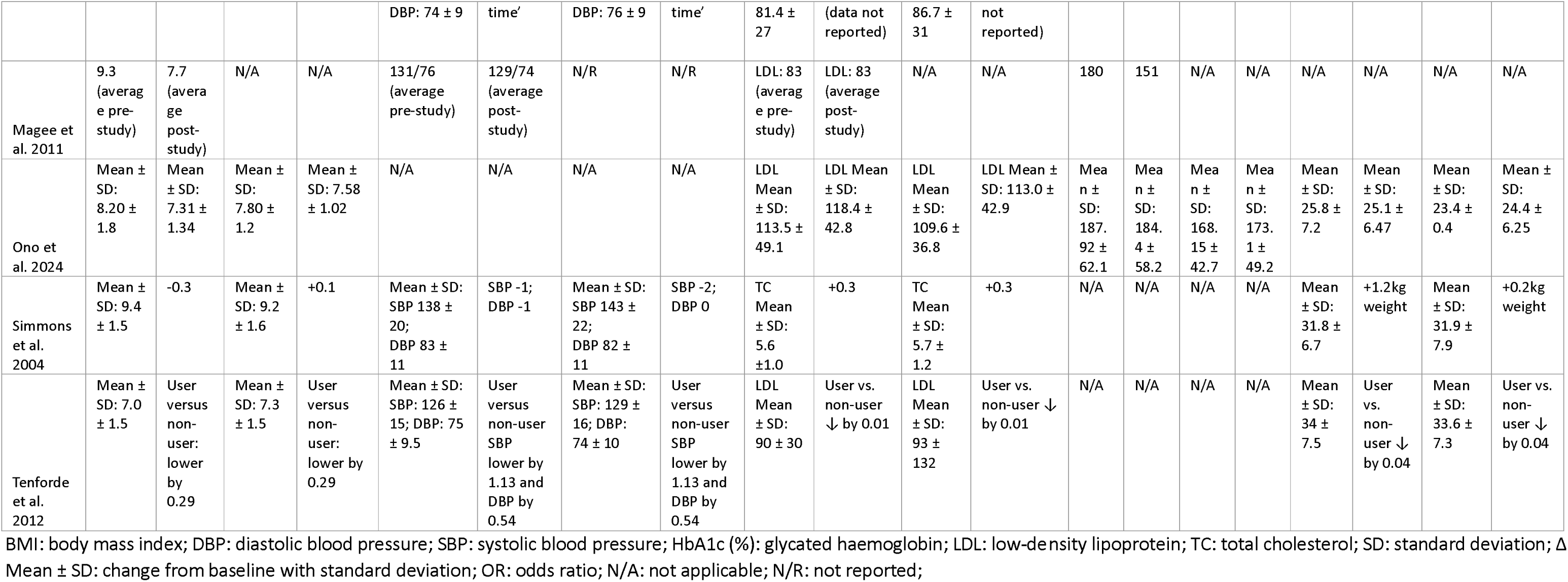
Overview of the Results for Primary Outcome.

**Table 3.** Thematic Synthesis of Patients’ Opinions and Perceptions.

| Theme (Subtheme) | Study | Key Findings |
| --- | --- | --- |
| **1. Usability** |  |  |
| <u>1.1 Barriers and Challenges</u> | Azizi<sup>4</sup> | - Unintuitive icons/units hindered use. |
|  | Fuji<sup>52</sup> | - Excessive data entry demands.<br>- Difficult to use |
|  | Fuji<sup>53</sup> | -Technical issues (e.g., internet access, hardware). |
| <u>1.2 Improved Functionality</u> | Azizi<sup>4</sup> | - Prepopulated data and color-coded alerts needed |
|  | Fuji<sup>52</sup> | - Creation of personalised functions<br>- Promote easy access |
| **2. Perceived Value** |  |  |
| <u>2.1 Self-Management and empowerment</u> | Dijkstra<sup>51</sup> | -Better control of diabetes management |
|  | Fuji<sup>53</sup> | -Useful for tracking trends |
|  | Simmons<sup>57</sup> | -No empowerment effect |
| <u>2.2 Knowledge</u> | Dijkstra<sup>51</sup> | -Increase in diabetes knowledge |
|  | Simmons | -No increase in knowledge |
| <u>2.3 Added Value</u> | Fuji<sup>52</sup> | -Non-personalized tools |
|  | Fuji<sup>53</sup> | -Lacked value |
| **3. Collaboration** |  |  |
| <u>3.1 Provider Engagement</u> | Fuji<sup>53</sup> | - Providers did not use PHR data |
|  | Dijkstra<sup>51</sup> | - Improved team coordination |
| <u>3.2 Patient Engagement</u> | Fuji<sup>52</sup> | -Low engagement |
|  | Fuji<sup>53</sup> |  |
| <u>3.2 Care Coordination</u> | Dijkstra<sup>51</sup> | -Better structure of care<br>-More shared of responsibilities |

#### 4.4.2 Results for Primary Outcome (Diabetes Health Outcomes)

Table 2 presents an overview of the results for primary outcomes, including HbA1C, blood pressure, cholesterol, blood glucose, and BMI.

#### Results for HbA1C

Eleven studies reported on the HbA1C outcome. One study^3^ did not report intervention or comparison baseline data. Two studies^74, 77^ reported the average data outcomes. One study^73^ reported results using the odds ratio. One study^80^ did not report result data for the follow-up period.

Figure 3 presents the results of the meta-analysis of five RCT studies^71, 72, 75, 76, 79^ evaluating mean difference (MD) and standard deviation (SD) in HbA1c (%). Moderate heterogeneity was observed among the included studies (I² = 68%); therefore, a fixed-effects model was employed. The pooled analysis demonstrated a statistically significant reduction in HbA1c in favour of the intervention group (MD −0.19; 95% CI −0.31 to −0.07; p = 0.001). GRADE appraisal identified this as low-certainty evidence, as the evidence was downgraded for high or unclear risk of bias across multiple domains and inconsistency among studies.

**Figure 3:**
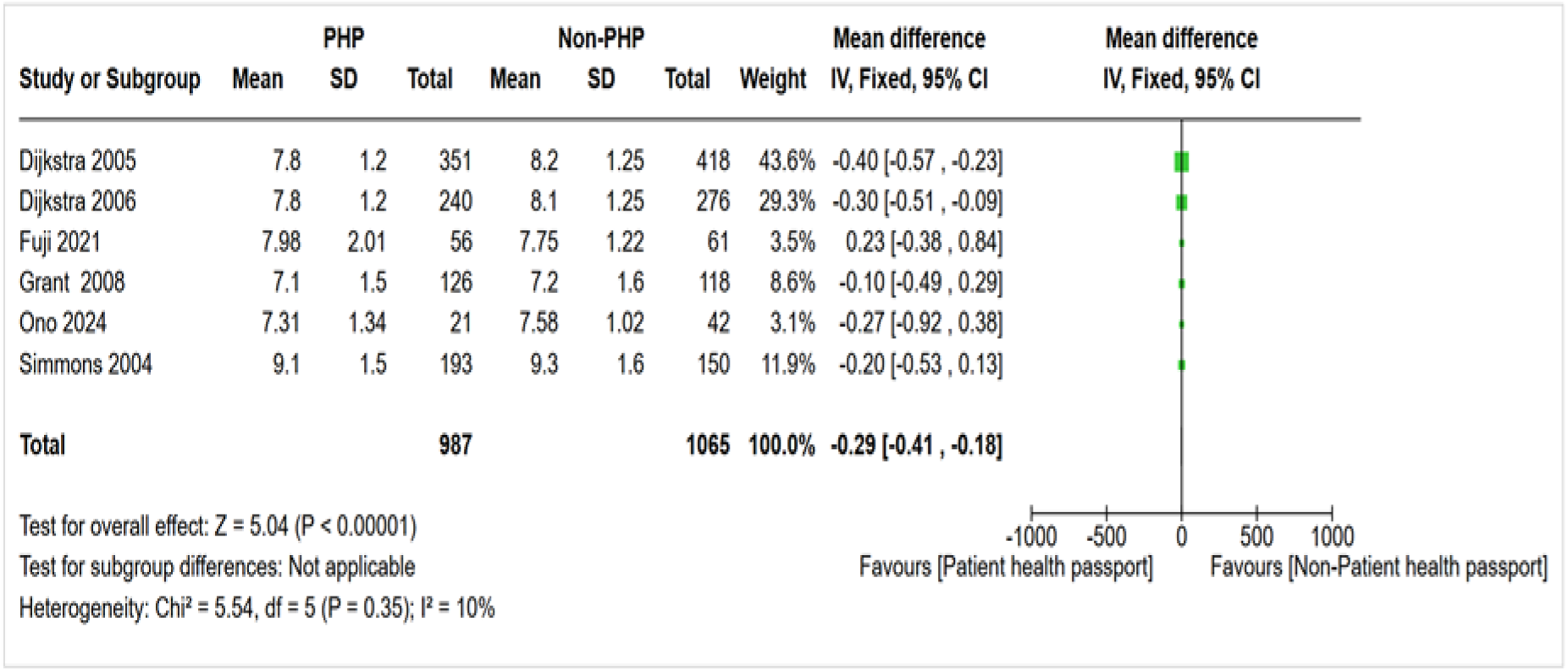
**Forest plot of the impact on the HbA1C (%)**

**Figure 4:**
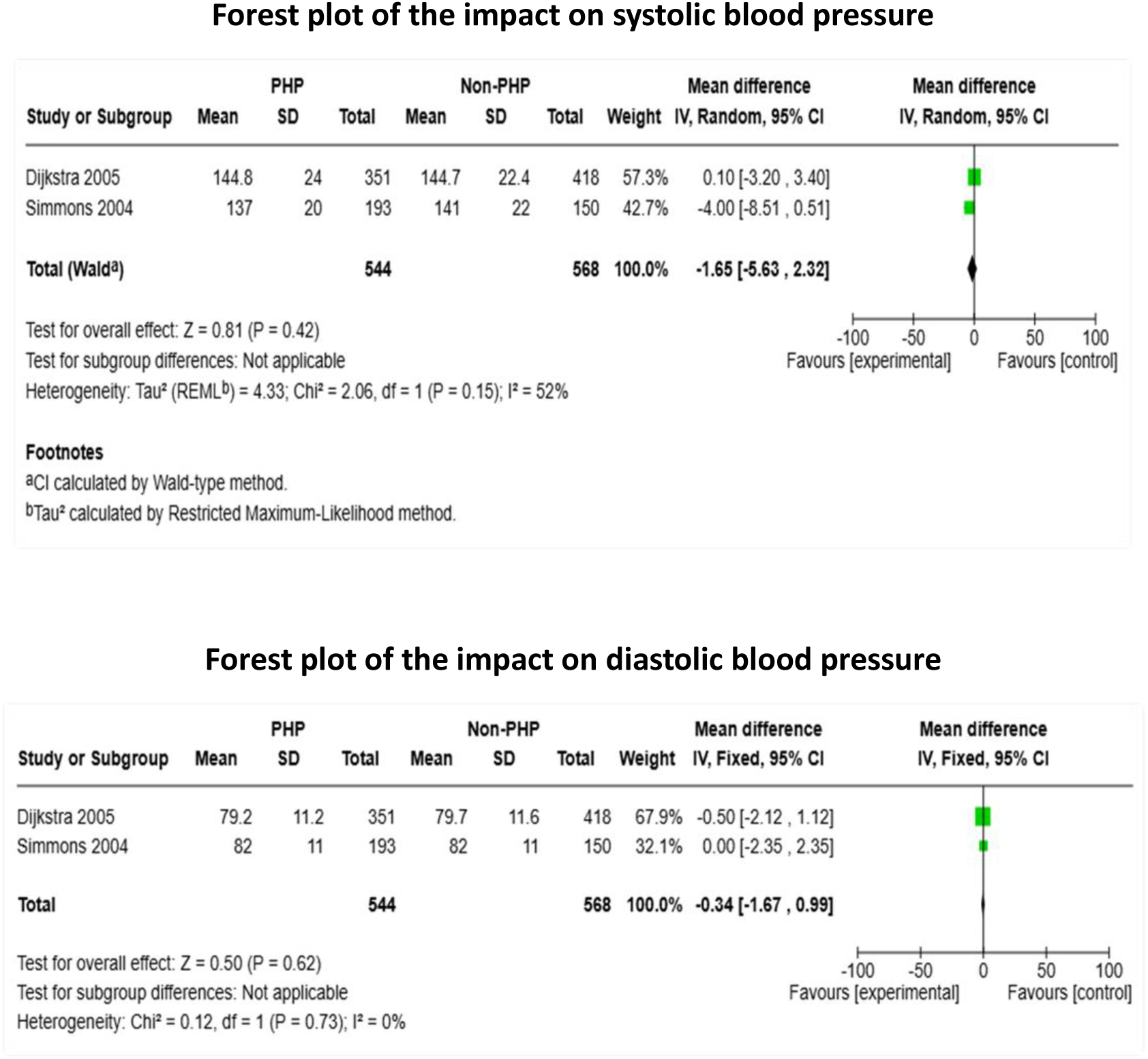
**a Forest plot of the impact on systolic blood pressure (mmHg); b: Forest plot of the impact on diastolic blood pressure (mmHg)**

For the remaining studies ^3, 73, 74, 77, 78, 80^ not included in the meta-analysis, a narrative synthesis of the outcome is provided. One study^3^ showed no MD in average HbA1c for both groups at study end (Mean ± SD: −0.2 ±0.1, both groups). A further study^73^ reported an increased odds of achieving the target HbA1c of < 8.5% in both study groups (Intervention: OR: 91.6 pre; OR: 92.8 post; Control: OR: 89.5 pre; OR: 88.7 post). Conversely, one study^74^ reported an increase in HbA1c of 0.32 (7.46 to 7.78) in the intervention group, from baseline to follow up, with no change noted in the control group. Magee^77^ reported an average HbA1c of 9.3 in the pre-test group, versus 7.7 in the post-test group. This indicates a decrease of 1.6 units from baseline to the end of the study. Tenforde^80^ stated that HbA1c was lower by 0.29 in the intervention group. Lastly, one study^78^ reported a non-significant reduction in HbA1c in favour of the intervention group (MD −0.27; 95% CI −0.92 to 0.38). The wide confidence intervals reflect the small sample size and limited statistical power of the study.

#### Results for Blood Pressure

Nine^3, 71–73, 75–77, 79, 80^ studies reported on the impact on BP. One study^3^ did not report specific data but presented a graph instead, and one study^75^ did not report follow-up data. One study^73^ reports results in odds ratios. Three studies^72, 76, 80^ did not report numeric data for the follow-up period (see Table 2).

Figure 4a presents the results of the meta-analysis of two studies^71, 79^ evaluating the MD (SD) in systolic blood pressure (SBP) (mmHg). Moderate heterogeneity was observed (*I*^2^=52%), so a random effects model was employed. The pooled analysis shows an MD of −1.65 (95% CI: −5.63 to 2.32; *p*= 0.42), indicating no statistically significant difference in SBP between the two study groups. GRADE appraisal identified this as very low certainty evidence, because it was downgraded twice for high or unclear risk of bias across multiple domains, once for heterogeneity and once for imprecision due to a wide confidence interval which crosses zero.

Figure 4b presents the results of the meta-analysis of two studies^71, 79^ evaluating the MD (SD) in Diastolic Blood Pressure (DBP) (mmHg). No significant heterogeneity was observed among the included studies (*I^2^*=0%), therefore, a fixed effect model was employed. The pooled analysis shows a MD of −0.34 (95% CI: −1.67 to 0.99; *p*= 0.62), indicating no statistically significant difference in DBP between the two study groups. GRADE appraisal identified this as very low certainty evidence, because it was downgraded twice for high or unclear risk of bias across multiple domains and once for imprecision due to a wide confidence interval which crosses zero. For the remaining studies not included in the meta-analysis and where data are available^73, 77, 80^ a narrative synthesis is provided. Dijkstra^73^ reported an increase in the OR of achieving BP targets in both groups, however, the control group showed better overall ORs at study end (Intervention: SBP<150 OR: 58.5 DBP <85 OR: 77.5; Control: SBP <150, OR: 59.3; DBP <85 OR: 78.2). Magee^77^ reported a reduction in average BP (from 131/76 mmHg to 129/74 mmHg) from pre-test to post-test. Tenforde^80^ reported that in the intervention group versus control group, the SBP lowered by 1.13 mmHg and DBP lowered by 0.54 mmHg.

#### Results for Cholesterol

Ten studies analysed cholesterol. Five studies^75–78, 80^ analysed LDL levels, four studies^71–73, 79^ analysed total cholesterol (TC) and one study^3^ analysed both LDL and TC. One study^73^ reports results as odds ratio. Three studies^72, 76, 80^ did not report numeric data for the follow-up period and one study ^75^ did not report follow-up data results (see table 2).

Figure 5 presents the results of the meta-analysis of two studies^71, 79^ evaluating the MD (SD) in TC (mmol/L). No significant heterogeneity was observed among the included studies (*I^2^*=0%), therefore a fixed effects model was employed. The pooled analysis shows an MD of −0.18 (95% CI: −0.29 to −0.06; p= 0.003), indicating a statistically significant difference in TC (mmol/L) in favour of the intervention group. GRADE appraisal identified this as low certainty evidence, because it was downgraded twice for high or unclear risk of bias across multiple domains.

**Figure 5:**
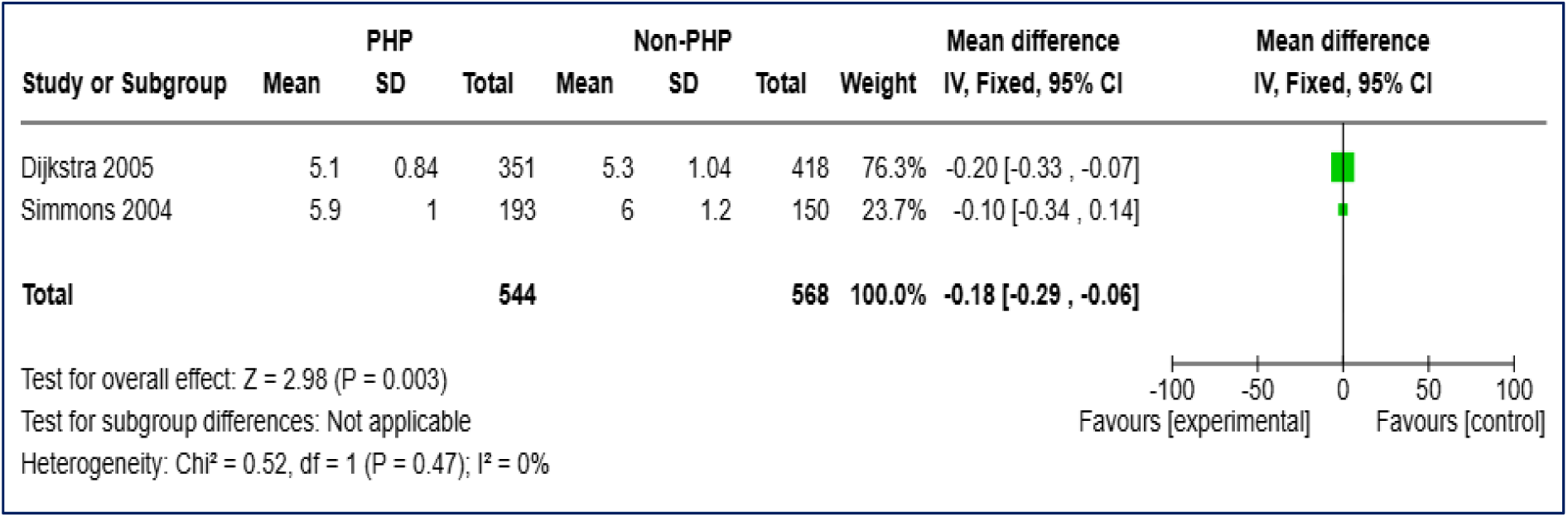
**Forest plot of the impact on total cholesterol (mmol/L)**

For the remaining studies not included in the meta-analysis, and where data are available^3, 72, 73, 76–78, 80^, a narrative synthesis is provided. Two studies^3, 80^ reported that cholesterol was reduced in the intervention group versus the control group. Dijkstra^73^ reported achieving improved odds of reaching TC targets (<5 mmol/L) in both groups. Two studies report no change in cholesterol^72, 77^ over the study period, whereas one study^76^ reported an improvement in cholesterol over time. Finally, in Ono^78^ there was an increase in mean LDL in the intervention group from baseline to follow up (MD: 4.90, 95% CI: −22.96 to 32.76). Similarly, there was an increase in mean LDL in the control group from baseline to follow up (MD: 3.40, 95% CI: −20.77 to 27.57).

#### Results for Blood Glucose

Four studies^74, 75, 77, 78^ analysed BG. Due to the nature of the reporting of the data, no meta-analysis was undertaken. Fuji^74^ showed an increase in the average BG in the intervention group (151.1 mg/dL to 156.5 mg/dL) and the control group (187 mg/dL to 205.9 mg/dL). In contrast, Magee^77^ showed a decrease from 180 mg/dL at baseline to 151 mg/dL post-intervention. In Ono^78^ there was a decrease in BG in the intervention group from baseline to follow up (MD: −3.52 mg/dL, 95% CI:-39.92 mg/dL to 32.88 mg/dL). In the control group, there was an increase in BG (MD: 4.95 mg/dL, 95% CI: −25.92 mg/dL to 35.82 mg/dL) One study^75^ did not report follow-up data results (see table 2).

#### Results for BMI

Five studies^72, 75, 78–80^ analysed BMI (see Table 2). Due to the nature of the reporting of the data, no meta-analysis was undertaken. Two studies^72, 75^ did not report follow-up data results. Simmons^79^ reported results as weight and not BMI, noting an increase of 1.2 kg of weight in the intervention group compared to an increase of 0.2kg in the comparison group. In Ono^78^ there was MD of-0.70 Kg/m^2^ [95% CI:-4.91 Kg/m^2^, 3.51 Kg/m^2^] in BMI in the intervention group, from baseline to follow-up. In the control group, there was a mean difference of 1.00 Kg/m^2^ (95% −1.68 kg/m^2^, 3.68 Kg/m^2^) from baseline to follow up, showing a greater reduction in BMI in the control group. Tenforde^80^ reported that BMI reduced in the intervention group versus the control group by 0.04 Kg/m^2^.

#### 4.5.2 Results for the Secondary Outcome Patient’s

##### Opinions and Perceptions

Table 3 reports a thematic synthesis of five studies^3, 73–75, 79^ exploring patients’ opinions and perceptions on the usage of PHR and patients’ passports. This analysis revealed three analytic themes: (1) usability, (2) perceived value and (3) collaboration, each with distinct subthemes. Usability emerged as a central theme due to frequent reports of design and technical challenges affecting PHR adoption. Within this theme, the subtheme *barriers and challenges* captured issues such as unintuitive interfaces^3^, excessive data entry demands, difficulty to use^74^ and technical limitations^75^. These barriers were consistently cited as reasons for low engagement by the patients. Conversely, the subtheme *improved functionality* highlighted user-driven recommendations, including prepopulated data^3^, color-coded alerts^3^ and mobile accessibility^74^, suggesting that improvements in these areas could increase usability and sustained use.

Perceived value reflected mixed patient experiences regarding the benefits of PHPp and PHRs. The subtheme *self-management and empowerment* demonstrated divergent findings, with some studies reporting improved control of diabetes management^73^ and tracking capabilities^75^, while others found no significant impact on self-care behaviours^79^. Similarly, *knowledge enhancement* produced conflicting results, with one study reporting increased disease knowledge^73^ and other^79^ observed no change. The subtheme *added value* further underscored variability, as some users viewed PHRs as redundant compared to existing tools^75^, emphasising the need for personalised features^74^ to enhance relevance.

Finally, collaboration emerged as a theme, with findings underscoring the role of provider and patient engagement. The subtheme *provider engagement* revealed that providers would disregard PHR data, relying instead on traditional methods, which diminished the tool’s utility^75^. In contrast, one study reported active provider involvement noted improved team care coordination^73^. The subtheme *patient engagement* highlighted low usage rates due to time constraints^74^ and poor usability^75^, while *care coordination* demonstrated that PHPp could facilitate shared decision-making and structured care^73^.

## 5. Discussion

The aim of this systematic review was to evaluate the impact of using a PHP on health outcomes among adults with diabetes, and 11 studies^3, 71–80^ met the inclusion criteria. The focus of the review was on HbA1c, BP, BG, and Cholesterol levels.

HbA1c was the only marker analysed across all the included studies, indicating its universal recognition as a key indicator of diabetes management effectiveness and reinforcing its central role in clinical guidelines for monitoring long-term glucose control^11, 12^. This emphasises the importance of including HbA1c tracking as a core feature in the PHPp and PHRs. The meta-analysis including five studies^71, 72, 75, 76, 79^ showed a statistically significant improvement in HbA1C in favour of the intervention group, but the evidence is of low certainty. A narrative analysis of three studies^73, 77, 80^, showed improvements in HbA1c and one study^78^ reported a non-significant reduction in HbA1c. Conversely, one study^3^ showed no difference in average HbA1c for both groups at study end and a further study^74^ reported an increase in HbA1c in the intervention group with no change noted in the control group. It is argued that measurement of HbA1c is essential in the management of individuals with diabetes^81^, in that is provides insights into how well diabetes is being controlled and managed. It is evident from the findings in this review that, the use of a PHP has a positive impact on HbA1c levels and as such, points to the usefulness of the PHP in the day-to-day care of patients with diabetes.

BP and cholesterol levels were also frequently reported, underscoring their relevance as comorbid risk factors in individuals with diabetes^16^. Nine ^3, 71–73, 75–77, 79, 80^ studies reported on the impact on BP. Results of the meta-analysis, including two studies^71, 79^, identified no statistically significant difference in either systolic or diastolic blood pressure among the study groups, with very low certainty evidence. Results from the narrative analysis of three studies showed mixed results, with one study^73^ reporting an increase in the OR of achieving BP targets in both groups, but the control group showed a better overall OR. A further study^77^ reported a reduction in average BP, and the final study^80^ reported a better reduction in BP in the intervention group. Ten studies analysed cholesterol ^3, 71–73, 75–80^. The results of the meta-analysis of two studies^71, 79^ indicated a statistically significant difference in TC (mmol/L) in favour of the intervention group, with low certainty evidence. Findings from the narrative synthesis of eight studies^3, 71–73, 76–80^ were conflicting. Two studies^72, 77^ reported no change in cholesterol levels, four studies^3, 73, 76, 80^ reported improvements over time, while two studies^78, 79^ observed increases in cholesterol in both study groups during the study period. The findings here suggest that the PHPs have a limited standalone impact on improving BP and cholesterol control. These findings may reflect the multifactorial nature of BP and lipid management, which often requires sustained behavioural change, medication adherence and clinical oversight^82^, factors not guaranteed by the implementation of the PHP alone. This highlights the need for PHP to be integrated into broader, system-level features to support sustained behavioural change^83^.

BG monitoring was underrepresented in the reviewed studies and was only reported in three studies^74, 77, 78^. This limited inclusion is notable given the importance of real-time glucose variability in diabetes self-management^84^. While HbA1c remains the gold standard marker for long-term glycaemic control, it fails to capture short-term fluctuations such as hyperglycaemia and hypoglycaemia, which are critical for timely clinical intervention and patient decision-making^13^. In this context, BG tracking could be more effectively supported by PHRs equipped with real-time feedback functionalities. The underreporting of BG may result from several factors, such as early-generation of PHR systems, which may lack integration with continuous glucose monitoring devices or manual input support, limiting their capacity to capture and respond to real-time data^83^. Findings of this review revealed only minor fluctuations in average glucose levels, with no significant differences between intervention and control groups, suggesting either limited intervention effectiveness or low user engagement with BG tracking tools.

BMI results were reported in only three studies^78–80^, also indicating a limited acknowledgment of weight management in diabetes care. Given the strong association between obesity and type 2 diabetes, this represents a gap in evaluation that may overlook a key factor in diabetes progression^85^. Evidence suggests that even a modest weight reduction of approximately 5% can lead to significant improvements in HbA1c, LDL, and TG levels, and increase HDL cholesterol^86^. Overall, the findings of this review regarding BMI were mixed and inconclusive and combined with the incomplete follow-up data and inconsistent reporting, limit the possibility of drawing firm conclusions. Studies also suggest that BMI change, like BP and lipid control, likely requires multi-component behavioural support beyond digital record access alone^78^.

For the secondary outcome, patients’ opinions and perceptions, analysis of the data identified three key themes: usability, perceived value and collaboration. These themes provide a nuanced understanding of both the facilitators and barriers to the usage of PHPp and PHRs in the management of diabetes, from the user perspective. Usability emerged as a foundational determinant of patient engagement while using PHRs^87^. Technical and design-related challenges were consistently identified as barriers to sustained use^3, 74, 75^. These findings are in line with previous digital health literature, which emphasises the importance of tailored design and intuitive interfaces in promoting user engagement and adherence^88, 89^. Further, evidence suggests that personalisation and contextual relevance are critical for driving sustained health behaviour change^74, 83^.

The theme of perceived value revealed mixed experiences with the impact of PHPp and PHRs on diabetes self-management and patients’ empowerment. Empowering patients to take an active role in managing their health is central to modern healthcare delivery, as it enhances treatment outcomes and supports long-term wellbeing^90^. Providing individuals with access to their health data and up-to-date information fosters awareness and encourages proactive disease management, ultimately improving consultation effectiveness and quality of life^91^. Evidence that using both PHPp and PHR can improve the quality of patient management^20^ with patients reporting improved ability to monitor and manage their condition^73, 75^. Simmons^79^ reported no changes in knowledge level using PHPp. Nonetheless, Dijkstra^73^ reported increased diabetes knowledge and control, highlighting the potential of PHPp.

Collaboration between patients and healthcare providers emerged as a key factor in the successful implementation of PHPp. The shift toward viewing patients as active partners rather than passive recipients of care reflects a broader transformation in healthcare, where sharing personal health data supports more collaborative, informed relationships^20, 92^. Findings in this review highlighted that when providers actively engaged with patient-entered data, PHPp facilitated shared decision-making and enhanced team-based care^73^. Conversely, providers who undermine their perceived value lead to discouraged patient use^75^. For an effective collaboration, both parties must recognize and engage with PHPp as integral to care delivery^73, 75^. However, patients’ engagement was often limited by practical barriers such as time constraints^74^ and poor usability^75^. Beyond promoting coordination, PHRs may strengthen communication among care teams and maintain continuity of care^20^.

With healthcare systems growing exponentially more complex, with larger groups of diverse healthcare professionals collectively working to deliver optimal patient care, there has been a wave of global interest in the potential of information technology to facilitate communication, reduce errors and ultimately improve quality of care^20^. This shift has benefited both patients and healthcare providers by enhancing communication and improving service delivery. However, and despite PHR benefits, there are still concerns regarding the security and privacy of personal health information. Research on PHRs has a strong focus on privacy, ownership and accessibility of patient data^93^. A study examined the privacy and security features of 52 free web-based PHR systems and found that they lacked detailed security measures and low compliance with standards and regulations^94^. Another study highlighted potential benefits of patients managing their own health data but noted a lack of studies detailing PHR capabilities and proven business cases to support their widespread use^93^. Moreover, literature on PHRs remains divided; some studies found that PHR usage increased healthcare interactions, such as visits, phone calls and hospitalizations, rather than reducing them^95^. Therefore, it is critical that future PHR development prioritise addressing concerns related to privacy, ownership, security standards and demonstrable value to both patients and healthcare systems.

### Limitations

The current SR has several limitations that should be considered when interpreting its findings. First, as this SR was restricted to the English language, we cannot rule out that additional relevant studies may have been excluded. The selected studies were limited by a relatively small sample size, which means that findings cannot be generalised^96^. Another significant limitation is the inconsistent and/or incomplete reporting of results across studies, diminishing the ability to form robust conclusions and to assess long-term effectiveness comprehensively. The studies’ follow-up durations limit the ability to evaluate long-term effects^97^ and with no follow-up exceeding 12 months, the sustainability of reported improvements remains uncertain^86^. Additionally, the evidence is geographically skewed toward high-income countries (USA, Netherlands, Iran, Japan and New Zealand). This is problematic for digital health interventions in low and middle-income countries which often face implementation barriers, such as limited internet access, low digital literacy and infrastructural challenges^98^. The study designs employed all showed a high risk of bias across multiple domains and GRADE appraisal, where relevant, identified the certainty of the evidence as low or very low. As a result, we cannot say with confidence what the impact of PHPs is on HbA1c, BP, BG and Cholesterol levels.

## 6. Conclusion

This SR evaluated the impact of patient health passports on health outcomes among adults with diabetes. Synthesis of the evidence from 11 studies showed mixed and inconclusive results. Findings suggest that PHP, regardless of its format, may support improvements in HbA1c but shows little to no improvement on BMI, BP, BG, and cholesterol levels. This may reflect the multifactorial nature of diabetes management, which cannot be guaranteed by the stand-alone benefits of a PHP. Patient opinion and perceptions revealed some benefits in its usability, perceived value, and collaboration. However, the data emphasises the importance of user-centred, tailored design and provider engagement for its full potential effectiveness. This SR is limited by high risk of bias in the included studies, a low or very low certainty of evidence and variability of study quality, small sample sizes, short follow-up periods and inconsistent outcome reporting. Therefore, findings must be interpreted with caution, suggesting a need for future high-quality research with larger sample sizes.

## Data Availability

No new datasets were generated or analysed during the current study. All data included in this systematic review were obtained from previously published studies, which are cited in the reference list.

## Acknowledgments

Killian Walsh, Information Specialist, provided valuable assistance with the search strategy.

